# Understanding RSV Resurgence Following COVID-19 in Ontario, Canada: Evaluating the Roles of Contact Patterns and Maternal Immunity

**DOI:** 10.64898/2026.08.28.26361657

**Authors:** Alyssa Parpia, James Wright, Nisha Thampi, Ali Gharouni, Tiffany Fitzpatrick

## Abstract

**Background:** Respiratory syncytial virus (RSV) remains a leading cause of hospitalization in infancy, with severe outcomes influenced by both contact patterns and passive immunity. Non-pharmaceutical interventions (NPIs) during the COVID-19 pandemic suppressed RSV circulation and reduced opportunities for maternal immune boosting, potentially altering protection among newborns. We evaluated whether incorporating time-varying maternal immunity improves the ability of an age-structured transmission model to predict post-pandemic RSV hospitalization patterns in infants.

**Methods:** We analyzed population-based RSV hospitalizations among Ontario (Canada) infants (<1 year) from July 2, 2017 to June 25, 2024, using linked administrative databases. A deterministic compartmental model across seven age classes was calibrated against pre-pandemic data using Latin Hypercube Sampling. We compared a model incorporating time-varying contact rates alone against a specification that additionally included time-varying maternal immunity.

**Results:** Both specifications accurately reproduced pre-pandemic seasonality and macro-level post-pandemic resurgence features. The constant maternal immunity model showed slightly better accuracy in capturing the 2021/22 peak compared to the time-varying maternal immunity specification. However, both qualitatively captured the continued near-absence of RSV and the observed peak was captured within the 95% credible intervals. While both models precisely captured the timing and overwhelming surge of admissions that occurred in 2022/23, they failed to capture the premature peak timing and magnitude in 2023/24.

**Conclusions:** Incorporating time-varying maternal immunity did not improve model accuracy post-pandemic. While maternal protection is essential for evaluating infant immunizations, population-level contact shifts primarily shaped post-pandemic RSV seasonality, indicating that models must account for these mechanisms of RSV transmission dynamics.

**Summary:** Using population-based Ontario hospitalization data, we evaluated whether incorporating time-varying maternal immunity improves an age-structured transmission model‘s ability to reproduce infant RSV disease patterns. While maternal immunity did impact post-pandemic RSV resurgences, we found that population-level contact pattern shifts were primarily responsible.

## Background

Respiratory syncytial virus (RSV) remains a leading cause of hospitalization in infancy, yet accurately modelling infant RSV disease is challenging because the determinants of severe infant outcomes are not limited to infant contact patterns or population-level transmission.(1) Infants, particularly in the first few months of life, rely on maternally-derived antibodies for protection against severe disease.(2) This creates a modelling challenge: while epidemic timing tends to be shaped by transmission in older children and the broader population, the observed hospitalization burden is concentrated in infants whose risk of infection is influenced by passive immunity.

The COVID-19 pandemic created a natural perturbation in RSV circulation that provides an opportunity to examine this issue.(3,4) Non-pharmaceutical interventions (NPIs), such as school closures and stay-at-home orders, suppressed RSV transmission and altered contact patterns. However, these interventions also reduced opportunities for recurrent RSV infections to boost the immune system of women of childbearing age.(3,5) A period of limited maternal immune boosting could reduce the proportion of infants born with optimal passive protection, changing infant hospitalization patterns driven primarily by transmission among older children and adult household members.(6) A better understanding of these mechanisms has direct relevance for informing new RSV prevention programs, including maternal vaccination and long-acting monoclonal antibodies, and for informing future immunization strategies that may target preschool- or school-aged children, as important drivers of transmission.(7)

Using RSV-related hospitalization data for all infants living in Ontario, Canada’s largest province, we examined changes in RSV seasonality following COVID-19 pandemic-related disruptions as a test case to evaluate whether incorporating time-varying maternal immunity improves the ability of an age-structured transmission model to reproduce RSV disease patterns in infants. We compared two model specifications: one incorporating time-varying age-specific contact rates alone, and one that additionally incorporates time-varying maternal immunity.

## Methods

### Population and Setting

All RSV-related hospitalizations occurring in Ontario among infants <1 year of age from July 2, 2017 to June 25, 2024 were identified from the Canadian Institute for Health Information’s Discharge Abstract Database (CIHI-DAD), a population-based source of hospital discharge abstracts linked to the Registered Persons Database (RPDB), a registry of Ontario residents. Individual-level databases were linked using unique encoded identifiers and analyzed at ICES.(8) The use of data in this project was authorized under section 45 of Ontario’s Personal Health Information Protection Act and exempt from research ethics board review.

RSV-related hospitalizations were defined as any admission with an RSV-related diagnostic code recorded anywhere on the discharge abstract using International Classification of Diseases, Tenth Revision (ICD-10) codes for RSV pneumonia (J12.1), acute bronchiolitis due to RSV (J21.0), acute bronchitis due to RSV (J20.5), or RSV as the cause of disease classified elsewhere (B97.4). These codes have been previously validated in this population and found to have both high specificity and sensitivity.(9) Weekly RSV-related admission rates were calculated using population-based denominators.

### Model Structure

We developed an age-structured, deterministic compartmental model of RSV transmission across the Ontario population. The model included compartments for maternally-derived immunity (M), primary susceptibility (S1), latent primary infection (E1), infectious primary infection (I1), temporary post-infection immunity (R), susceptibility to subsequent infection (S2), latent subsequent infection (E2), and infectious subsequent infection (I2). Primary and subsequent infections were modeled separately to allow for differences in infectiousness and in the probability of hospitalization following repeat RSV infection.

The population was divided into seven age classes: <1, 1 to <2, 2 to <5, 5 to <20, 20 to <50, 50 to <65, and ≥65 years. Individuals aged between compartments using a cohort-aging approach. Births entered the infant age class and deaths occurred from the oldest age group, consistent with a stable population assumption. Age-specific mixing was based on the POLYMOD contact matrix (United Kingdom) as implemented in the R socialmixr package and symmetrized for use in the deterministic transmission model.

The model was implemented in R using the deSolve package and simulated using daily time steps. A schematic diagram of the model is provided in **Appendix A.1 (Figure S1)**.

### Model Parameters

Model parameters were either fixed using literature-informed values or estimated through model fitting **(Table 1)**. We assumed a stable Ontario population of approximately 14.2 million people, a life expectancy of 82 years, and a birth rate equal to the death rate. The latency period, infectious period, duration of natural immunity, duration of maternally-derived immunity, baseline transmission coefficient, seasonal forcing amplitude, and seasonal phase shift were estimated during calibration. Subsequent infections were assumed to be less infectious than primary infections, with infectiousness reduced by 25%. Seasonal variation in RSV transmission was represented using a cosine forcing function. Plausible ranges for fitted parameters were identified from the literature and sampled using Latin hypercube sampling.

**Table 1.** Epidemiological, Seasonal, and Disease Severity Parameters Used in the RSV Transmission Model.

| Parameter | Meaning<sup>a</sup> | Value (95% CrI)<sup>b</sup> | Source<sup>c</sup> | References |
| --- | --- | --- | --- | --- |
| **Epidemiological and Immunological** |  |  |  |  |
| $\epsilon$ | Latency period (days) | 2.8 (2.0 – 5.5) | Fitted [2 – 6] | (16,23) |
| $\gamma$ | Infectious period (days) | 8.5 (7.1 – 11.9) | Fitted [7 – 21] | (24,25) |
| $\rho$ | Natural immunity duration (days) | 158.1 (101.4 – 261.5) | Fitted [100 – 280] | (23,26) |
| $\rho_M$ | Maternal immunity duration (days) | 104.0 (92.6 – 118.5) | Fitted [90 – 120] | (6,23) |
| $c$ | Reduced infectiousness of secondary infections | 75% | Literature | (22,23,26) |
| **Seasonal Transmission** |  |  |  |  |
| $\beta_0$ | Baseline transmission rate | 0.02 (0.01 – 0.02) | Fitted [0.01 – 0.02] | (22,27) |
| $\beta_1$ | Seasonal amplitude | 0.30 (0.18 – 0.40) | Fitted [0.10 – 0.40] | (22,27) |
| $\Phi$ | Peak phase shift (years) | 0.40 (0.36 – 0.45) | Fitted [0.35 – 0.50] | (22,27) |
| **Age-Specific Hospitalization Probability** |  |  |  |  |
| $h_1$ | Primary infection | <1 yo: 3.00%<br>1 - <2 yo: 0.96%<br>2 - 4 yo: 0.70%<br>5 - 19 yo: 0.05%<br>20 - 49 yo: 0.05%<br>50 - 64 yo: 0.10%<br>65+ yo: 0.50% | Literature | (1,28,29) |
| $h_2$ | Subsequent infection | 40% x $h_1$ | Literature | (23,26) |

a Model parameters governing transitions between epidemiologic states were implemented as rates (per day) within the differential equations framework. For interpretability, durations are reported as the inverse of these rates (e.g., latency period = $1/\epsilon$ ).
b Median and 95% credible interval (CrI) shown for fitted parameters.
c Sampling range [minimum – maximum] shown for fitted parameters.

### Time-varying contact rates during the COVID-19 period

To account for changes in social mixing during the COVID-19 pandemic and subsequent recovery period, we applied time-varying, age-specific multipliers to the baseline contact matrix. These multipliers reflected the timing and relative intensity of major pandemic-related changes in Ontario, including school closures, stay-at-home orders, phased reopening, return to in-person schooling and childcare, and the post-restriction resurgence period.(10,11) A multiplier of 1 represents pre-pandemic contact intensity; values below 1 represent reductions in contacts during periods of NPIs, while values above 1 represent periods of above-baseline mixing or enhanced pediatric contact intensity during the rebound period. Further details, including the full set of calendar periods and age-specific contact multipliers, are provided in **Appendix A.2**.

### Time-varying maternal immunity

Because the primary outcome used for calibration and validation was RSV-related hospitalization among infants, and because infants are uniquely dependent on passively-acquired maternal antibodies, we extended the model to allow the proportion of newborns entering the maternally-immune compartment to vary over time. This time-varying maternal immunity component was intended to capture reduced maternal RSV immune boosting during periods of suppressed RSV circulation, followed by partial recovery as RSV recirculated. The maternal immunity function was aligned with the pandemic/NPI chronology and scaled to reflect reduced opportunities for natural immune boosting during periods of low RSV transmission and reduced social mixing. In the base post-pandemic specification, maternal protection among newborns was assumed to be at pre-pandemic levels before the disruption of RSV circulation, markedly reduced following the missed 2020/21 RSV season, partially restored during the 2021/22 recirculation period, and returned to baseline following the intense 2022/23 resurgence.(12,13)

Infants in the M compartment transitioned to primary susceptibility at the fitted rate of waning maternally-derived immunity. Thus, time-varying maternal immunity affected the size of the infant population initially protected from primary RSV infection and severe disease, while the duration of protection was estimated through model fitting. The duration of protection once an infant entered the maternally protected compartment was unchanged. Further details are provided in **Appendix A.3**.

### Hospitalization probabilities

Incident RSV infections were converted to RSV-related hospital admissions by applying literature-informed age- and infection-history-specific hospitalization probabilities. The probability of hospitalization was highest for primary infections in infancy and reduced for subsequent infections to reflect lower severity after prior exposure. Weekly model-predicted admissions among infants were expressed as rates per 1,000 infants to match the observed data.

### Model fitting and uncertainty

The model was initialized with a multi-year burn-in period and calibrated to observed weekly RSV-related admission rates among Ontario infants during the pre-pandemic period (July 2, 2017 through March 17, 2020, inclusive). We used Latin hypercube sampling to draw 10,000 parameter sets across literature-based ranges for latency period, infectious period, duration of natural immunity, duration of maternally-derived immunity, baseline transmission coefficient, seasonal forcing amplitude, and seasonal phase shift. For each parameter set, weekly predicted infant admissions were compared with observed weekly admissions using a joint Poisson likelihood based on weekly incident and cumulative admissions. The top 1% of parameter sets, ranked by likelihood, were retained as the best-fitting ensemble. Parameter medians and 95% credible intervals (CrIs) were calculated from this ensemble.

### Pandemic and post-pandemic model evaluation

Following pre-pandemic calibration, we used the pandemic and post-pandemic period as a model-validation test case to evaluate the added value of explicitly accounting for maternal immunity for predicting infant RSV disease. Two specifications were compared, incorporating 1) time-varying age-specific contact rates reflecting changes in social mixing during the COVID-19 pandemic and recovery period but assumed constant maternally-derived immunity; and 2) an additional time-varying maternal immunity component representing reduced passive protection among newborns following prolonged suppression of RSV circulation and gradual recovery after RSV re-emergence.

For each specification, predicted weekly RSV hospitalization rates among infants aged <1 year were compared with observed rates. Model performance was assessed using three complementary outcomes: weekly admission trajectories over time, epidemic peak timing, and epidemic peak magnitude. This approach allowed us to evaluate not only whether the model reproduced total burden, but also whether it captured the timing and intensity of seasonal epidemics.

Respiratory seasons were defined from July 1 through June 30. Peak timing was defined as the epidemiologic week with the highest weekly RSV hospitalization rate within a season, and peak magnitude was defined as the maximum weekly hospitalization rate per 1,000 infants. For each season from 2017/18 through 2023/24, observed peak timing and magnitude were compared with model-estimated medians and 95% CrIs from the top 1% of best-fitting parameter sets. Weekly epidemic curves were visualized using observed data alongside median model estimates and 95% CrIs. Peak timing distributions were summarized using boxplots across retained parameter sets.

All analyses were performed in R Version 4.6.1. Supplementary methodological details, including the full set of ordinary differential equations **(Appendix A.4)**, are provided in **Appendix A**.

## Results

### Pre-Pandemic Model Calibration (2017–2020)

Prior to evaluating pandemic-era disruptions, the age-structured transmission model was calibrated to weekly infant RSV hospitalization rates. The top 1% best-fitting parameter ensemble **(Table 1; Supplementary Figure 1)** demonstrated strong fidelity to baseline epidemiological dynamics. Across the 2017/18, 2018/19, and 2019/20 respiratory seasons, the model median run closely tracked annual winter seasonality, peak timing, and peak admission magnitudes. Aside from some minor deviations in 2019/20—which was impacted by the COVID-19 pandemic, observed weekly rates consistently fell within the model-estimated 95% CrI **(Figure 1)**.

**Figure 1.**
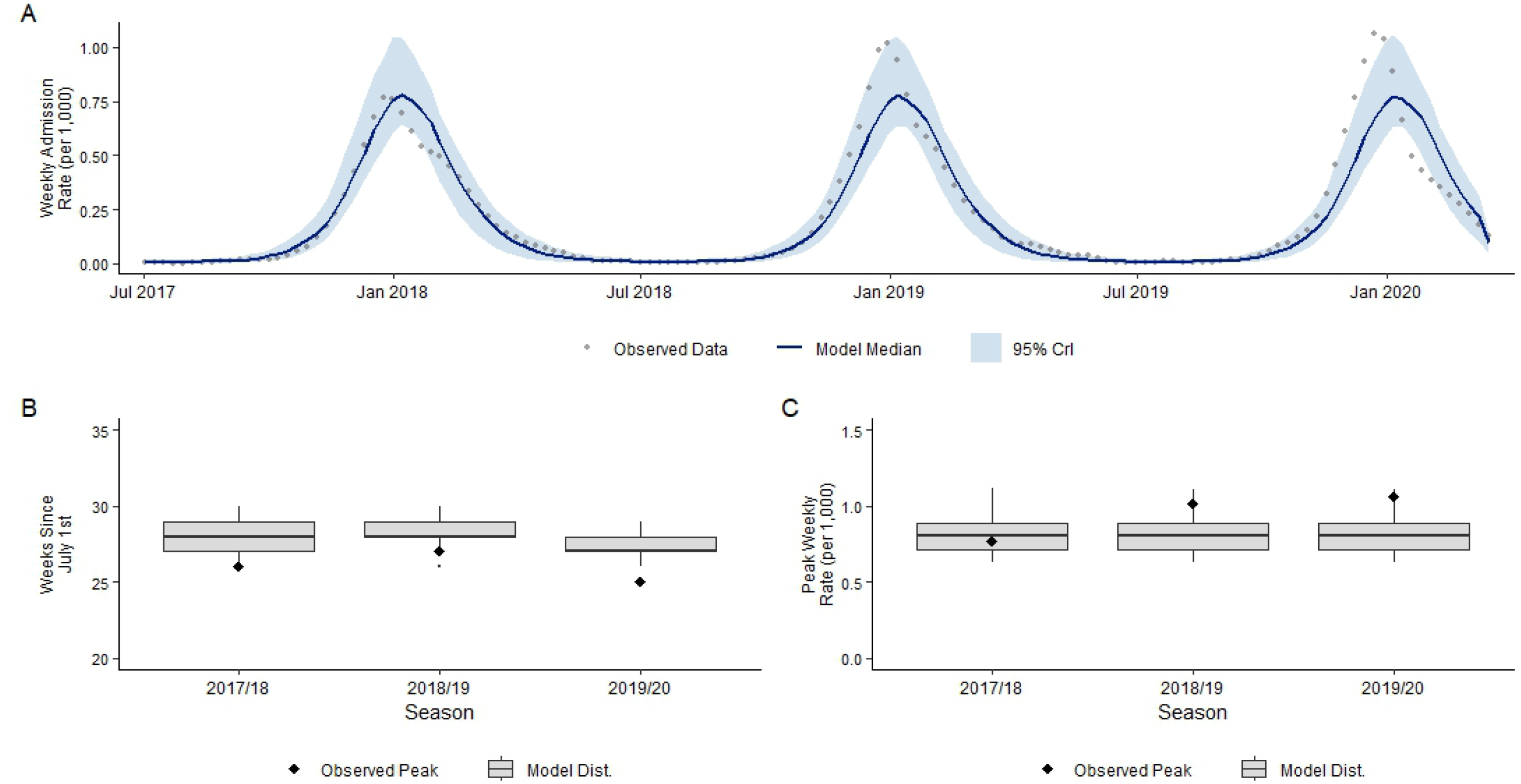
Pre-pandemic calibration model performance among infants (<1 year); July 2017 – March 2020. **Panel A** illustrates weekly observed (grey circles) and model-predicted (median trajectory; dark blue line, with 95% CrI ribbon) RSV hospitalization rates per 1,000 infants during the pre-pandemic calibration window (July 2, 2017 – March 17, 2020). **Panel B** compares model-estimated (gray boxplots showing median, interquartile range, and full distribution across top-performing Latin Hypercube Sampling runs) and observed (diamonds) peak epidemic timing, measured in weeks since July 1st, across the 2017/18, 2018/19, and 2019/20 respiratory seasons. **Panel C** compares model-estimated (gray boxplots) and observed (diamonds) peak weekly admission rates per 1,000 infants across the same three pre-pandemic respiratory seasons.

### Maternal Immunity Specification Comparison

Overall, the model specification utilizing time-varying contact rates with constant maternal immunity yielded admission trajectories that were quite similar to the extended specification incorporating dynamic maternal antibody decay **(Figure 2)**.

**Figure 2.**
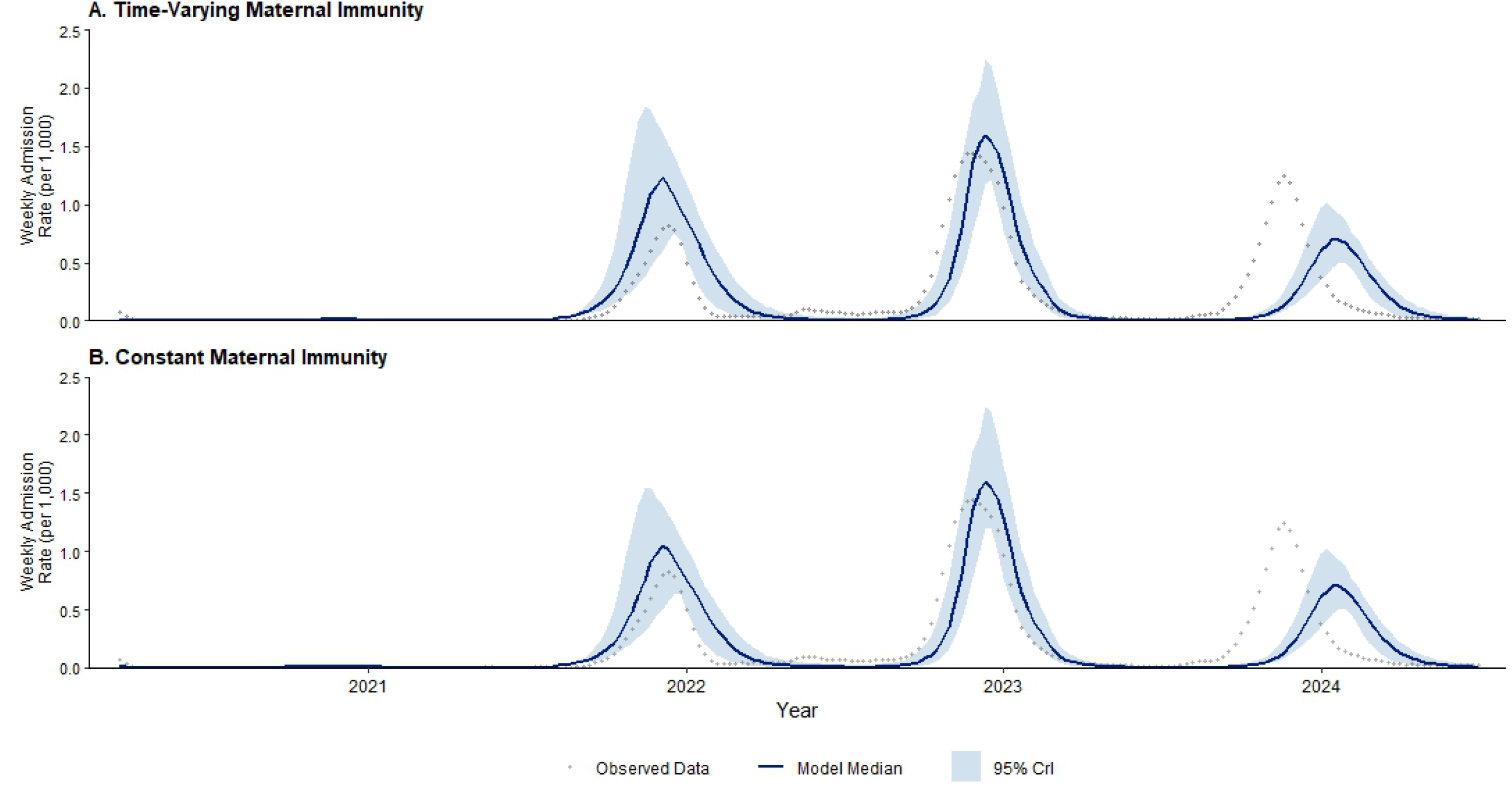
Observed and predicted weekly RSV admission rates among Ontario infants (<1 year of age), March 2020 to June 2024, across model specifications. **Panel A** illustrates the structural model specification incorporating time-varying maternal immunity decay. **Panel B** illustrates the baseline comparative specification maintaining a uniform maternal immunity fraction. In both panels, charcoal circles represent empirical weekly observed data. The solid dark blue line represents the continuous ensemble model median run, bounded by the light blue shaded ribbon detailing the 95% credible interval (CrI).

However, some notable differences emerged when analyzing discrete seasonal peak properties **(Figure 3; Supplementary Table 1)**. For the 2021/22 respiratory season, both models slightly overestimated the observed peak magnitude (0.006 per 1,000); albeit both models accurately suggested a near-absence of RSV admissions that season (estimated peak, time-varying immunity: 0.014 [95% CrI: 0.013-0.015]; constant immunity: 0.012 [95% CrI: 0.011-0.012]. The observed peak was also slightly overestimated by both models during the 2021/22 season, with this difference being slightly more apparent among the time-varying approach; however, both successfully captured the observed peak within their respective 95% CrIs [observed: 0.822; time-varying: 1.280 [95% CrI: 0.775-1.886]; constant: 1.095 [95% CrI: 0.669-1.597]. Similarly, both models captured successfully the size of the subsequent substantial 2022/23 surge; observed: 1.440; time-varying: 1.704 [95% CrI: 1.339–2.246]; constant: 1.711 [95% CrI: 1.352–2.249]. Notably, both models failed to adequately capture the magnitude of the 2023/24 season and, instead, incorrectly estimated a lower-than-observed peak; observed: 1.244; time-varying: 0.741 [95% CrI:0.525–1.015]; constant: 0.744 [95% CrI: 0.527–1.021].

**Figure 3.**
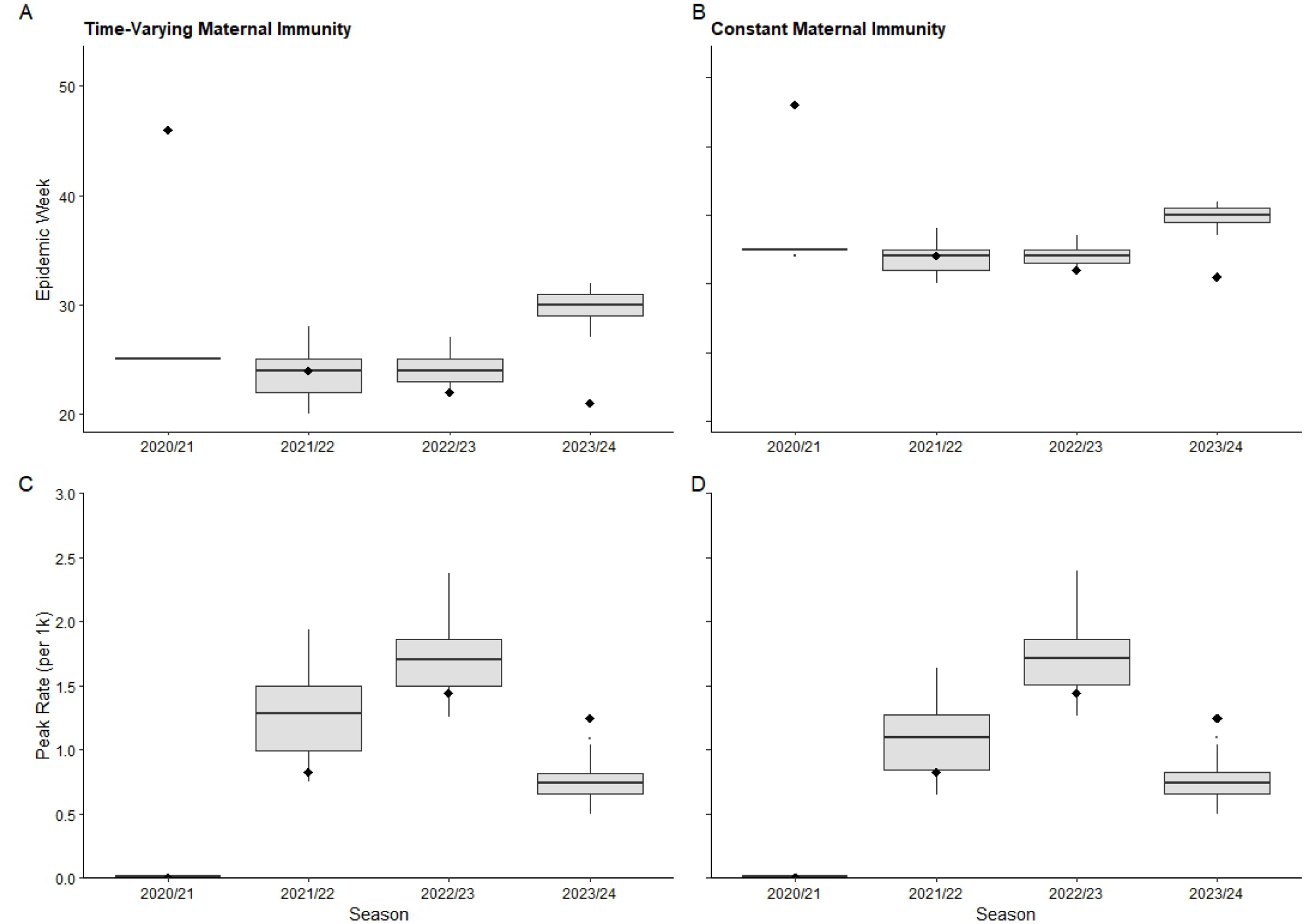
Predicted and observed RSV seasonal epidemic timing and magnitude, 2020/21 – 2023/24, across model specifications. The top row contrasts predicted epidemic peak timing (weeks since July 1st) between the proposed model with time-varying maternal immunity (**Panel A**) and the baseline model with constant maternal immunity (**Panel B**). The bottom row contrasts the peak weekly admission rate per 1,000 infants between the time-varying (**Panel C**) and constant (**Panel D**) specifications. Gray boxplots display the LHS model distribution (top 1% parameter sets), while black diamonds indicate the observed values.

Similar observations were made with respect to the ability of both models to accurately capture the observed seasonal peak timing **(Figure 3; Supplementary Table 1)**. Neither model was able to accurately capture the out-of-season, albeit substantially attenuated, return of RSV in summer 2020/21; instead, both suggested RSV admissions would peak in during mid-December. In contrast, both accurately estimated that the 2021/22 season would peak the week of December 12. While both models suggested the 2022/23 season would peak a few weeks later than observed, both accurately captured the observed peak week in their respective CrIs; observed: November 27; time-varying: December 11 [95% CrI: November 27 to January 1]; constant: December 11 [95% CrI: November 27 to January 1]. Both approaches failed to capture the earlier-than-expected peak in 2023/24 and instead suggested the peak would occur weeks later than typically seen pre- and post-pandemic; observed: November 19; time-varying: January 21 [95% CrI: December 31 to February 4]; constant: January 21 [95% CrI: December 31 to February 4].

While not validated against observed data, both models also suggested virtually indistinguishable post-pandemic RSV-related hospitalization trajectories among older cohorts aged 1+ years **(Supplementary Figure 2)**.

## Discussion

In this study, we utilized the epidemiological disruptions following the COVID-19 pandemic as a natural stress test to evaluate the structural role of maternal immunity in influencing infant RSV transmission. Early theoretical frameworks anticipated that COVID-19-related NPIs would dramatically suppress endemic viral circulation, generating an expanded susceptible cohort— often conceptualized as an “immunity debt”—that would fuel atypical resurgences upon societal reopening.(3) Evaluating model trajectories across this post-resurgence period revealed a high degree of structural convergence between constant and time-varying maternal immunity specifications. Both frameworks successfully reproduced the macro-level features of the RSV resurgence. Crucially, this capability was most likely attributable to our explicit incorporation of time-varying contact rates, suggesting that epidemic intensity was primarily driven by temporal behavioral shifts rather than shifts in passive immunity alone. This aligns with classical age-stratified transmission modeling frameworks demonstrating that seasonal RSV resurgences are fundamentally governed by shifting mixing patterns across age cohorts,(14) as well as modern analyses emphasizing the necessity of robust age-structured mixing for capturing post-pandemic RSV dynamics.(15)

When assessing discrete outbreak properties; however, the baseline specification utilizing constant maternal immunity demonstrated superior predictive accuracy for the empirical peak magnitude of the initial 2021/22 epidemic wave. The extended model incorporating dynamic maternal antibody decay slightly overestimated this initial peak, though the observed provincial peak remained well within the ensemble‘s 95% CrI. This alignment implies that population-level passive protection may possess greater structural resilience than parsimonious step-reduction functions assume, or that adult antibody titers decayed more slowly during the pandemic period than expected. Alternatively, this minor overestimation may reflect unobserved behavioral countermeasures, such as parental cocooning or targeted physical distancing during early reopening phases, which temporarily insulated infants. Empirical serological investigations conducted in British Columbia, Canada provide unique evidence documenting that marked temporal reductions in RSV antibody titers did occur among women of childbearing age and infants following prolonged NPI-induced viral suppression, with antibody levels subsequently recovering alongside viral recirculation.(12,13) In all, our findings should be interpreted as evidence that this simple population-level maternal immunity specification did not improve model performance, rather than evidence that maternal immunity is an unimportant driver of RSV dynamics.

Notably, both model formulations failed to adequately capture the atypical dynamics of the 2023/24 season; thus, neither maternal immunity nor contact patterns modifications alone are sufficient to explain post-pandemic shifts in RSV dynamics. Given the widespread susceptibility depletion and elevated maternal boosting following the severe 2022/23 wave, a prominent delay in subsequent circulation would have been expected; consequently, one might reasonably hypothesize that the subsequent season would occur earlier than typical and be of lower magnitude. Instead, RSV re-emerged unexpectedly early and peaked at a magnitude similar to pre-pandemic levels. This discrepancy underscores that community-wide viral circulation is governed by transmission drivers operating independently of the localized mechanisms which dictate infant RSV hospital admissions. For example, preschool- and school-aged children act as primary engines of community-level amplification due to high-contact environments (e.g., daycares and schools) but remain underrepresented in severe-disease surveillance data.(16–18) Furthermore, this miscalibration may have been compounded by unmeasured shifts in healthcare-seeking behavior, changes in multi-pathogen diagnostic testing thresholds and practices following the pandemic,(19) or viral interference dynamics with co-circulating respiratory pathogens (such as influenza and SARS-CoV-2)(20,21) that can alter clinical presentation and hospitalization rates independently of underlying RSV transmission intensity.

Despite these nuances, explicit representation of passive immunity remains essential for accurate contemporary RSV modeling. In particular, the need for accurate models is especially important for understanding and evaluating changes in RSV burden within the current evolving public health landscape—marked by the widespread rollout of maternal immunizations and long-acting monoclonal antibodies. While parsimonious models using fixed background parameters may be able to successfully replicate historical RSV data, they lack the mechanistic architecture required to simulate the direct effects of these biological interventions (e.g., maternal immunity). Moreover, robust modelling frameworks and intervention strategies must account for broader RSV transmission vectors. While interventions targeted solely at infants may successfully reduce severe clinical outcomes locally, they minimally impact community-wide transmission dynamics. Conversely, immunization programs targeting school-aged children could potentially yield substantial indirect benefits for infants by mitigating household introduction routes in multi-child homes. These findings carry critical implications for public health agencies deploying novel RSV prevention programs, such as long-acting monoclonal antibodies (e.g., nirsevimab) and maternal immunizations. Epidemic timing and community-wide transmission demonstrate substantial spatio-temporal variations and are fundamentally governed by broader transmission networks rather than infant-specific susceptibility alone, thus modelling frameworks must transcend the single-age cohort boundaries that have commonly been used (i.e., models exclusively considering transmission among infants). Crucially, policy makers and program planners implementing these programs and evaluating their population-level impacts must couple local clinical endpoints (e.g.., hospitalizations) with dynamic multi-age surveillance systems to accurately anticipate indirect effects and shift-timing of epidemic peaks; as these data are largely lacking in most judications, investments will be needed to gather more robust surveillance, capturing multiple disease severity endpoints across the age continuum.

Despite its many strengths, our study has some limitations. Although robust population-based data were used, the model was calibrated exclusively against infant hospitalization data, capturing only severe outcomes rather than mild community infections; however, this was partly mitigated through use of established, literature-based disease severity scaling parameters.(22) Moreover, time-varying contact multipliers served as simplified representations of macro-level pandemic policies and behaviors (e.g., masking, social distancing) without distinguishing household from community contacts; similarly, the maternal immunity function approximated population-level processes rather than directly measured changes in antibody titers (such as maternally derived immunity). Furthermore, the framework did not account for stochastic viral introductions; instead, RSV reintroduction was approximated using a deterministic seeding function. Alternative seeding assumptions may have influenced the timing of epidemic resurgence. The model also did not account for spatial heterogeneity, viral interference, or temporal shifts in healthcare-seeking behavior. Notwithstanding these limitations, the post-pandemic period offered a rigorous testbed for understanding RSV modeling mechanisms, highlighting that improving future predictive capacity requires enhanced surveillance of transmission dynamics across the broader age continuum—particularly among school-aged children—alongside refined data on and mechanistic representations of passive immunity.

## Conclusions

Using Ontario as a test case, we found that incorporating a simple time-varying maternal immunity extension did not improve model-based estimation of post-pandemic RSV hospitalization patterns among infants. While maternally derived protection is an important dynamic determinant of infant RSV disease and should be represented explicitly in models used to evaluate maternal vaccination, monoclonal antibody programs, and other infant RSV prevention strategies, these findings suggest that population-based changes in contact patterns may have played a more important role in shaping the observed post-COVID-19 resurgence pattern in RSV seasonality. However, the inability of both model specifications to capture the 2023/24 season also suggests that infant protection alone cannot explain RSV epidemic timing and highlights other drivers likely played a role in the RSV resurgence. In order to accurately estimate the potential direct and indirect impacts of new and emerging RSV immunization programs, a more robust understanding of the mechanisms driving RSV transmission across the age continuum is needed.

## Acknowledgements

This study uses data at ICES, which is funded by an annual grant from the Ontario Ministries of Health (MOH) and Long-Term Care (MLTC). Parts of this material are based on information compiled and provided by the Canadian Institute for Health Information (CIHI) and Ontario MOH. The analyses, conclusions, opinions, and statements expressed herein are solely those of the authors and do not reflect those of the data providers. No endorsement by ICES, the Ontario MOH or MLTC, its partners, or the Province of Ontario is intended or should be inferred.

## Author Contributions

Acquired funding: TF, AG, NT. Conceived and designed the study: TF, AG, NT. Analyzed the data: TF, AP, JW. Contributed analysis tools: TF. Provided clinical interpretation of the data: NT. Managed the study: TF. Contributed to drafting the manuscript: TF, AP. Reviewed and revised the manuscript: TF, AP, JW, AG, NT. All authors have read and approve the final manuscript.

## Data Sharing Statement

The data set from this study is held securely in coded form at ICES. While legal data sharing agreements between ICES and data providers (e.g., health care organizations and government) prohibit ICES from making the data set publicly available, access may be granted to those who meet pre-specified criteria for confidential access, available at www.ices.on.ca/DAS. The full data set creation plan and underlying analytic code are available from the authors upon request, understanding that the computer programs may rely upon coding templates or macros that are unique to ICES and are therefore either inaccessible or may require modification.

---

**Supplementary Figure 1.**
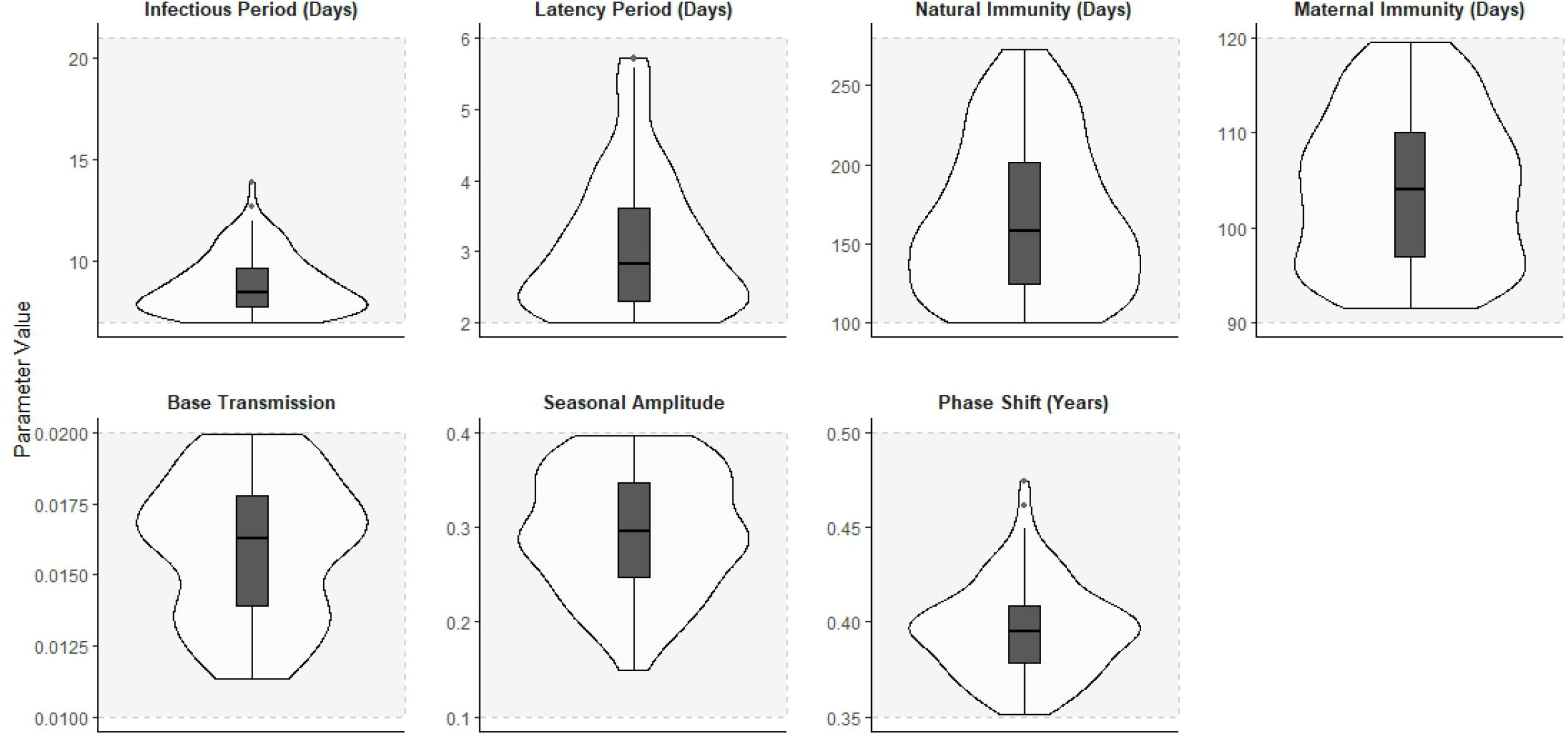
Parameter distributions of the top 1% best-fitting Latin Hypercube Sampling (LHS) iterations for the RSV transmission model. Violin plots illustrate the marginal parameter distributions derived from the top 1% best-fitting parameter sets generated via Latin Hypercube Sampling (LHS). Within each violin, the internal box plot displays the median (solid central horizontal line) and the interquartile range (shaded box), with vertical whiskers indicating data spread. Parameters evaluated include the infectious period (days), latency period (days), duration of natural immunity (days), duration of maternal immunity (days), base transmission rate, seasonal amplitude, and phase shift (years).

**Supplementary Figure 2.**
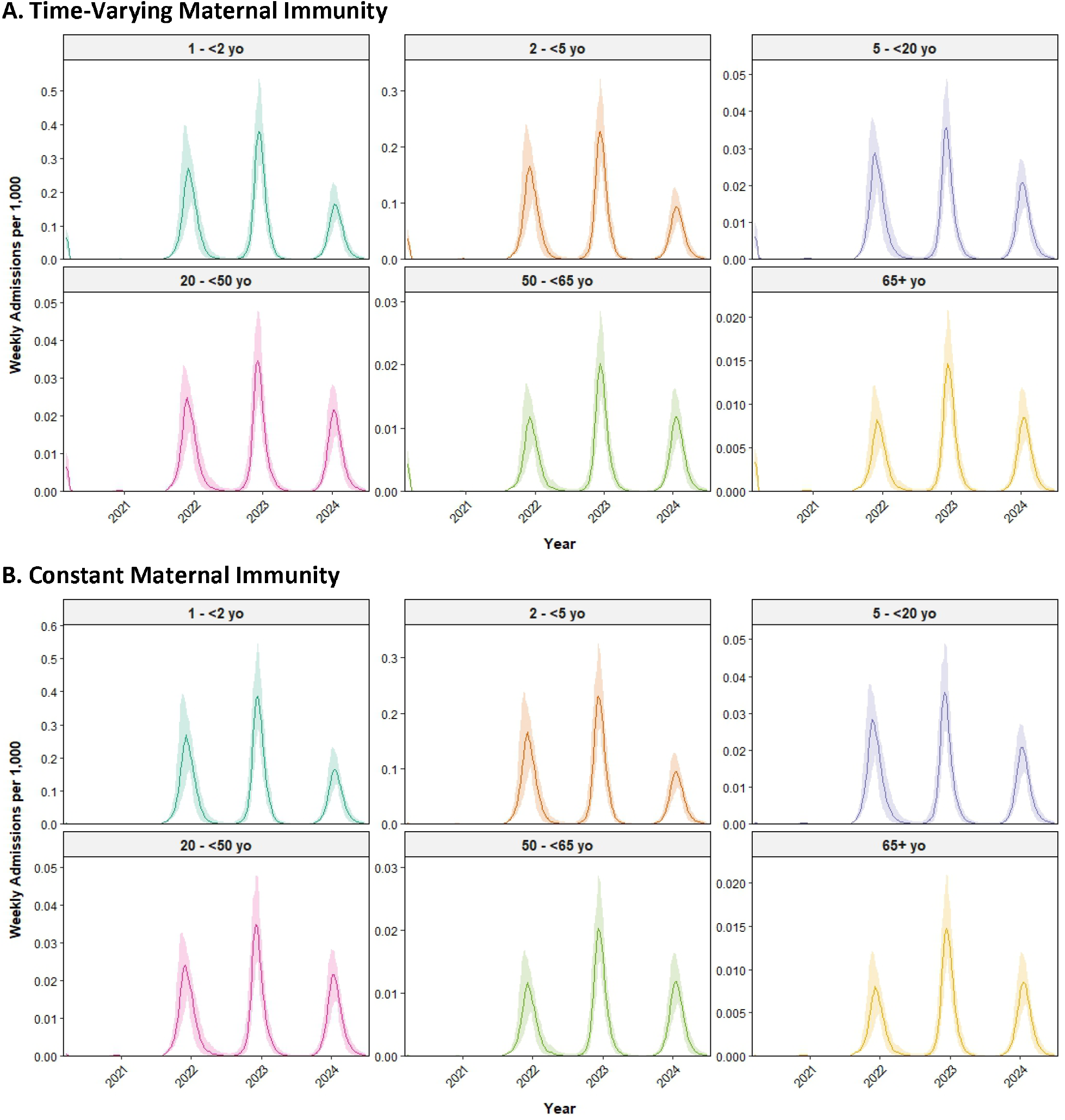
Model-projected weekly RSV hospitalization rates across older age cohorts (1+ years) comparing time-varying and constant maternal immunity specification; 2020–2024. Post-pandemic trajectory projections of weekly RSV hospital admissions per 1,000 individuals across six age strata (1–<2 years, 2–<5 years, 5–<20 years, 20–<50 years, 50–<65 years, and 65+ years) from March 2020 through June 2024. Solid lines represent the median trajectory and shaded ribbons denote the 95% credible intervals (CrI) derived from the top 1% best-fitting Latin Hypercube Sampling parameter sets.

**Supplementary Table 1.** Seasonal peak timing and rate of infant RSV hospitalizations 2020/21-2023/24: comparison of observed data with time-varying (TV) and constant maternal immunity model-based estimates.

| Season | Peak Rate<sup>a</sup> (95% CrI) |  |  | Peak Date (95% CrI) |  |  |
| --- | --- | --- | --- | --- | --- | --- |
|  | Observed | TV | Constant | Observed | TV | Constant |
| **2020/21** | 0.006 | 0.014<br>(0.013–0.015) | 0.012<br>(0.011–0.012) | 2021-05-16 | 2020-12-20<br>(2020-12-20 to 2020-12-20) | 2020-12-20<br>(2020-12-13 to 2020-12-20) |
| **2021/22** | 0.822 | 1.280<br>(0.775–1.886) | 1.095<br>(0.669–1.597) | 2021-12-12 | 2021-12-12<br>(2021-11-14 to 2022-01-02) | 2021-12-12<br>(2021-11-14 to 2022-01-02) |
| **2022/23** | 1.440 | 1.704<br>(1.339–2.246) | 1.711<br>(1.352–2.249) | 2022-11-27 | 2022-12-11<br>(2022-11-27 to 2023-01-01) | 2022-12-11<br>(2022-11-27 to 2023-01-01) |
| **2023/24** | 1.244 | 0.741<br>(0.525–1.015) | 0.744<br>(0.527–1.021) | 2023-11-19 | 2024-01-21<br>(2023-12-31 to 2024-02-04) | 2024-01-21<br>(2023-12-31 to 2024-02-04) |

a. Peak rates are expressed as weekly hospital admissions per 1,000 infants <1 year of age).
Abbreviations: TV, time-varying maternal immunity model.

## Appendix A Supplemental Methods

### A.1 Model Schematic and Structure

The RSV transmission dynamics were modeled using a deterministic, age-structured compartmental framework. A schematic diagram illustrating the model flow is provided in Figure S1. The model is structured into seven age classes, with individuals aging between compartments using a cohort-aging approach.

Newborns enter the model through a ‘Birth’ node and are routed into either the maternally protected compartment (M, light blue) or the naive susceptible compartment (S1, light blue) based on the time-varying maternal immunity function. The primary infection pathway (S1 E1 I1) leads to a recovered state (R, grey). Following the waning of natural immunity, individuals transition into the secondary susceptibility compartment (S2, light blue), which proceeds through subsequent latent (E2, pink) and infectious (I2, red) states before returning to the recovered state. Dashed lines indicate the probability of RSV-related hospital admission.

**Figure S1.**
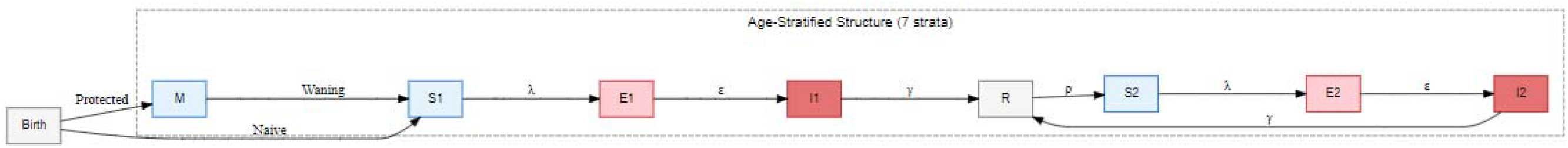
Schematic Representation of the Age-Stratified RSV Transmission Model. Boxes represent epidemiological compartments; arrows denote rates of transition. The model includes primary (S1-E1-I1) and secondary (S2-E2-I2) infection pathways, with a birth node routing newborns to either the maternally immune (M) or fully susceptible (S1) classes.

### A.2 Age-Stratified Time-Varying Social Contact Modifiers

To approximate the heterogeneous impact of COVID-19 NPIs and subsequent behavioral rebound on RSV transmission, we applied time-varying scalar modifiers to the age-stratified contact matrix. Multipliers represent the estimated net change in contact intensity relative to pre-pandemic baseline patterns, derived from provincial school operation status and Google Community Mobility indices.(10) Because our model uses a fixed contact matrix (based on pre-pandemic data), the multiplier acts as a pragmatic calibration parameter intended to capture the combined effects of pandemic-era changes in contact opportunities, behaviour, and transmission potential. The temporal dynamics of these modifiers across the study period are visualized in Figure S2, with further rationale for specific dates and values provided in Tables S2 and S3.

**Figure S2.**
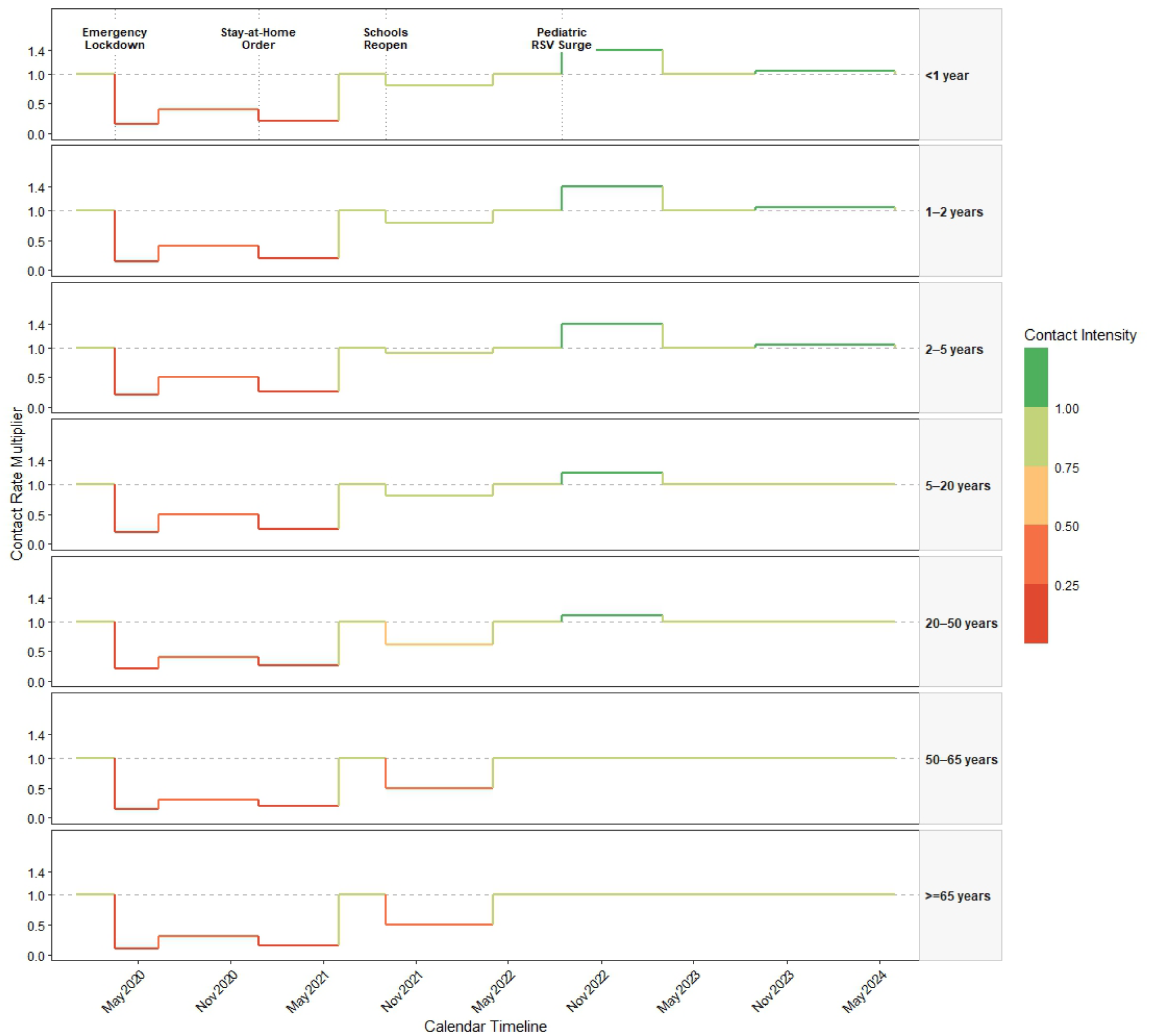
Time-Varying Contact Modifiers by Age Cohort (2020–2024). Step functions represent scalar modifiers applied to the contact matrix. Multipliers <1.0 indicate contact reduction due to NPIs.

**Table S1.** Justification of Time-Varying Social Contact Modifiers.

| **Date Range** | **Primary Policy / Mobility Driver** | **Justification of Multiplier** |
| --- | --- | --- |
| **Mar 2020 – Jun 2020** | **Strict Lockdown:** State of emergency, full school closures, and stay-at-home mandates. | **Contact Reduction:** Modifiers (0.15–0.20) reflect the ~80% reduction in retail, transit, and workplace mobility observed in Ontario Google Mobility datasets. |
| **Jun 2020 – Dec 2020** | **Tiered Reopening:** Schools partially open with reduced density; businesses operating under strict capacity limits. | **Intermediate Restriction:** Modifiers (0.30–0.50) represent a partial return to social activity constrained by physical distancing and masking mandates. |
| **Dec 2020 – Jun 2021** | **Winter Shutdown:** Re-imposition of province-wide lockdown following holiday surge. | **Contact Reduction:** Modifiers (0.15–0.25) return to near-initial lockdown levels to account for school closures and cessation of non-essential services. |
| **Sep 2021 – Apr 2022** | **Full School Return:** Return to full-time, in-person learning for all grades; removal of social distance requirements. | **Synchronization Effect:** Modifiers (0.70–1.10) account for the simultaneous return of two "immunologically naive" cohorts to high-density environments. |
| **Aug 2022 – Mar 2023** | **Behavioral Normalization:** Complete cessation of mask mandates; full return to pre-pandemic social mixing patterns. | **Intensity Scaling:** Multipliers (1.00–1.40) represent full contact return combined with the accumulated population-level susceptibility ("immunity debt") from previous years. |
| **Sep 2023 – Jun 2024** | **Endemic Equilibrium:** Established routine contact patterns; population susceptibility normalized. | **Baseline Normalization:** Modifiers (1.00–1.05) represent an endemic state where social contact has stabilized and "catch-up" effects have subsided. |

**Table S2.** Age-Specific Contact Rate Modifiers by Pandemic Era.

| **Date Period** | **<1 yo** | **1 - <2 yo** | **2 - 4 yo** | **5 - 19 yo** | **20 - 49 yo** | **50 - 64 yo** | **65+ yo** |
| --- | --- | --- | --- | --- | --- | --- | --- |
| **Baseline** | 1.00 | 1.00 | 1.00 | 1.00 | 1.00 | 1.00 | 1.00 |
| **Mar 2020 – Jun 2020** | 0.15 | 0.15 | 0.20 | 0.20 | 0.20 | 0.15 | 0.10 |
| **Jun 2020 – Dec 2020** | 0.40 | 0.40 | 0.50 | 0.50 | 0.40 | 0.30 | 0.30 |
| **Dec 2020 – Jun 2021** | 0.20 | 0.20 | 0.25 | 0.25 | 0.25 | 0.20 | 0.15 |
| **Sep 2021 – Apr 2022** | 1.10 | 1.10 | 1.10 | 1.00 | 0.80 | 0.70 | 0.70 |
| **Aug 2022 – Mar 2023** | 1.40 | 1.40 | 1.40 | 1.20 | 1.10 | 1.00 | 1.00 |
| **Sep 2023 – Jun 2024** | 1.05 | 1.05 | 1.05 | 1.00 | 1.00 | 1.00 | 1.00 |

### A.3 Time-Varying Maternal Immunity Function

To account for the immunological disruption caused by COVID-19-related non-pharmaceutical interventions (NPIs), we incorporated a dynamic function for maternal antibody protection. Literature suggests that population-level RSV immunity in adults is maintained through frequent, asymptomatic re-exposure in the community. Given the observed collapse of RSV circulation in Ontario from 2020 through mid-2021, we hypothesized this boosting effect was suspended, leading to a depletion of maternal antibody titers (IgG) transfer to newborns.

To mechanistically represent the reduction and subsequent re-priming of population-level maternal RSV antibody transfer, we applied a dynamic maternal protection fraction (*f*) to the birth-routing equation. This fraction represents the proportion of newborns entering the maternally protected compartment (M) versus the fully susceptible compartment (S1).

The maternal protection fraction was modeled as a time-varying parameter, with values assigned to represent an acute depletion phase (*f* =0.15) during the period of stringent NPIs, followed by a recovery phase (*f* =0.65) as community viral circulation resumed. These parameters serve as a mechanistic proxy for the decay and subsequent re-priming of population-level maternal immunity in the absence of longitudinal seroprevalence data.

The timeline of the maternal protection fraction was calibrated to reflect the observed progression of the COVID-19 pandemic and the subsequent recovery of RSV circulation in Ontario (Table S1). The timeline assumes a transition from a pre-pandemic baseline to a period of significant immunological suppression, followed by a phased recovery as community transmission patterns normalized.

**Table S3.**
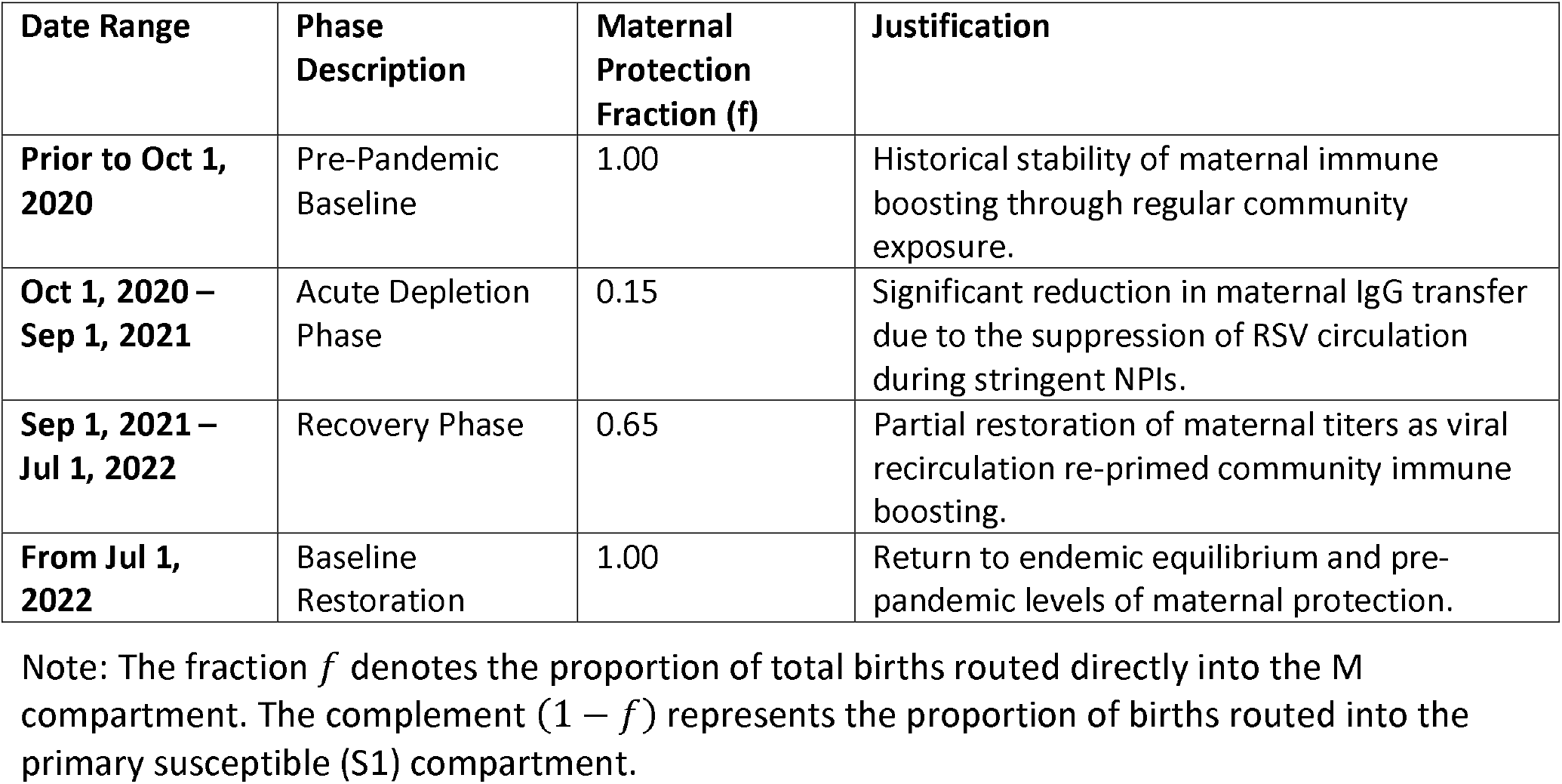
Time-Varying Maternal Protection Fraction Specifications.

| Date Range | Phase Description | Maternal Protection Fraction ($f$) | Justification |
| --- | --- | --- | --- |
| Prior to Oct 1, 2020 | Pre-Pandemic Baseline | 1.00 | Historical stability of maternal immune boosting through regular community exposure. |
| Oct 1, 2020 – Sep 1, 2021 | Acute Depletion Phase | 0.15 | Significant reduction in maternal IgG transfer due to the suppression of RSV circulation during stringent NPIs. |
| Sep 1, 2021 – Jul 1, 2022 | Recovery Phase | 0.65 | Partial restoration of maternal titers as viral recirculation re-primed community immune boosting. |
| From Jul 1, 2022 | Baseline Restoration | 1.00 | Return to endemic equilibrium and pre-pandemic levels of maternal protection. |

Note: The fraction $f$ denotes the proportion of total births routed directly into the M compartment. The complement ( $1 - f$ ) represents the proportion of births routed into the primary susceptible (S1) compartment.

### A.4 System Ordinary Differential Equations (ODEs)

The deterministic transmission dynamics are governed by the system of differential equations below. For each age stratum *a ∈* {1,…,7} the rate of change across the primary infection pathway (*M* → *S*_1_ → *E*_1_ → *I*_1_ → *R*) is defined as:

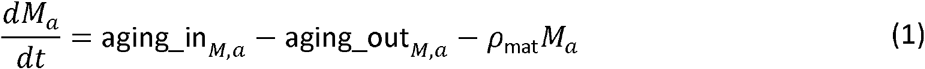

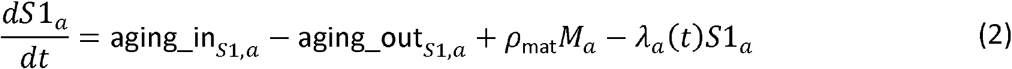

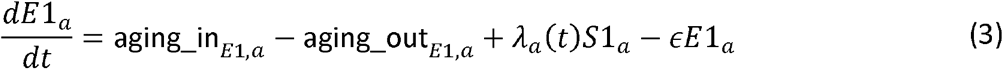

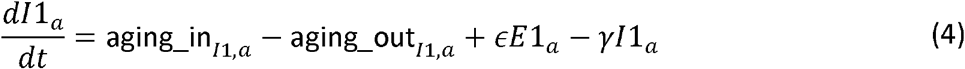

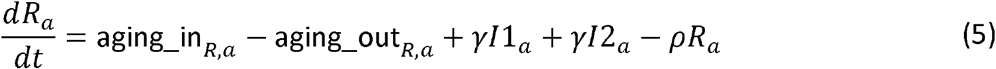

Following the loss of natural temporary immunity (*p*), individuals transition into the secondary susceptibility pathway (*S*_2_ → *E*_2_ → *I*_2_ → *R*):

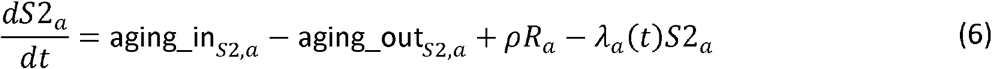

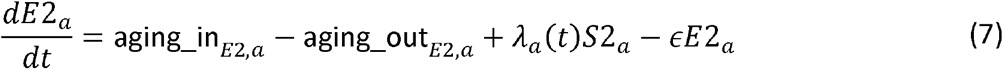

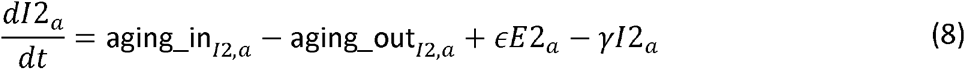

#### Force of Infection & Seasonal Forcing

The age-specific force of infection*λ*_*a*_(*t*), represents the instantaneous rate at which susceptible individuals in age group acquire RSV infection (i.e., new infections per susceptible individual per unit time, i.e., days). It incorporates the age-stratified contact matrix (*C*_*a,j*_), time-varying contact modifiers (*D*_*a*_(*t*)), reduced shedding during secondary infections (*c*), seasonal forcing (*q*_seasonal_), and a deterministic importation (seeding) term representing exogenous introduction of RSV into the modeled population:

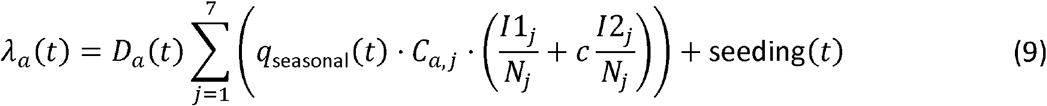

Specifically, to permit re-establishment of transmission following the suppression of RSV circulation during the COVID-19 pandemic, a low-level deterministic seeding term was added to the force of infection. Seeding is assigned a value of 0.0001 during periods of low seasonal transmission and 0.01 during periods of active seasonal transmission. To facilitate the observed resurgence following relaxation of public health restrictions, a larger seeding value (0.02) is applied from August 1 through October 31, 2021.

Seasonal variation in transmission is modeled via an annual cosine forcing function parameterized by baseline probability *q*_1_, amplitude *q*_2_, and phase shift Ф

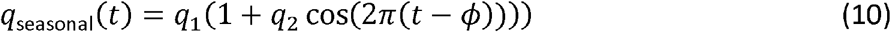

#### Birth Routing Mechanism

Births enter exclusively into the first age stratum (*a* = 1) To maintain a constant population baseline size, total births balance the total cohort mortality exit rate (*B*). Neonates are dynamically assigned to either the maternally immune (*M*_1_) or primary susceptible (*S*1_1_) compartment based on the maternal protection fraction *f*(t);

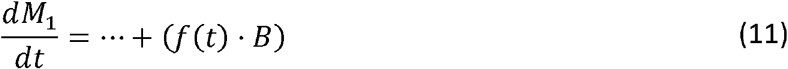

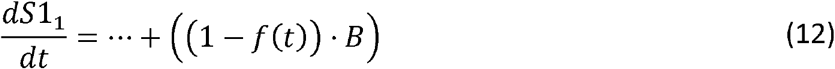

#### Cumulative Incidence Tracking

To evaluate hospital admission rates without numerical artifact from discrete prevalence steps, the system accumulates primary (*INC*1_*a*_) and secondary (*INC*2_*a*_) incident infections continuously:

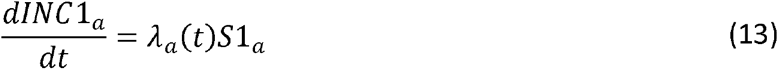

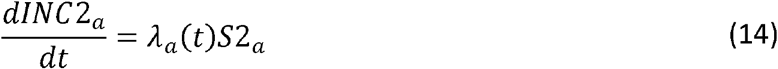

Daily hospital admissions for age group *a* are computed post-simulation by taking discrete daily differences (Δ) of these cumulative pools and scaling by age-specific primary (*h*_1,*a*_) and secondary (*h*_2,*a*_) hospitalization probabilities:

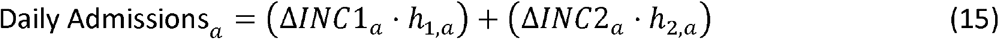

